# Chest X-Ray Has Poor Sensitivity and Prognostic Significance in COVID-19: A Propensity Matched Database Study

**DOI:** 10.1101/2020.07.07.20147934

**Authors:** Aditya Borakati, Adrian Perera, James Johnson, Tara Sood

**Author notes:** **Correspondence:** Dr Aditya Borakati, Academic Foundation Doctor, Emergency Department, Royal Free Hospital, London, UK NW3 2QG. **Author contribution (CRediT) statement: Aditya Borakati:** Conceptualization, Methodology, Validation, Formal Analysis, Investigation, Writing – Original Draft, Writing – Review & Editing, Visualization, Project Administration **Adrian Perera:** Conceptualization, Methodology, Investigation, Writing- Review & Editing, Supervision, Project Administration **James Johnson:** Investigation **Tara Sood:** Conceptualization, Methodology, Writing – Review & Editing, Supervision, Project Administration Aditya Borakati is the overall guarantor of this work. **Statistical review** The statistical methods in this manuscript and associated code have been reviewed by Dr Federico Ricciardi of the Department of Statistical Science at University College London and confirmed as robust and accurate. **Ethical approval** This study was registered with the local institutional review board as a service evaluation using anonymised data only. No formal ethics committee review was required. **Declarations of Interests** The authors have no relevant conflicts of interest to declare. All authors have completed the <u>Unified Competing Interest form</u> (available on request from the corresponding author) and declare: no support from any organisation for the submitted work; no financial relationships with any organisations that might have an interest in the submitted work in the previous three years, no other relationships or activities that could appear to have influenced the submitted work. **Transparency declaration** The lead author (AB) affirms that this manuscript is an honest, accurate, and transparent account of the study being reported; that no important aspects of the study have been omitted; and that any discrepancies from the study as planned (and, if relevant, registered) have been explained. **Copyright** The Corresponding Author has the right to grant on behalf of all authors and does grant on behalf of all authors, a worldwide licence to the Publishers and its licensees in perpetuity, in all forms, formats and media (whether known now or created in the future), to i) publish, reproduce, distribute, display and store the Contribution, ii) translate the Contribution into other languages, create adaptations, reprints, include within collections and create summaries, extracts and/or, abstracts of the Contribution, iii) create any other derivative work(s) based on the Contribution, iv) to exploit all subsidiary rights in the Contribution, v) the inclusion of electronic links from the Contribution to third party material where-ever it may be located; and, vi) licence any third party to do any or all of the above.

## Abstract

**Objectives:** To identify the diagnostic accuracy of common imaging modalities, chest X-ray (CXR) and computed tomography (CT) for diagnosis of COVID-19 in the general emergency population in the UK and to find the association between imaging features and outcomes in these patients.

**Design:** Retrospective analysis of electronic patient records

**Setting:** Tertiary academic health science centre and designated centre for high consequence infectious diseases in London, UK.

**Participants:** 1,198 patients who attended the emergency department with paired RT-PCR swabs for SARS-CoV 2 and CXR between 16^th^ March and 16^th^ April 2020

**Main outcome measures:** Sensitivity and specificity of CXR and CT for diagnosis of COVID-19 using the British Society of Thoracic Imaging reporting templates. Reference standard was any reverse transcriptase polymerase chain reaction (RT-PCR) positive naso-oropharyngeal swab within 30 days of attendance. Odds ratios of CXR in association with vital signs, laboratory values and 30-day outcomes were calculated.

**Results:** Sensitivity and specificity of CXR for COVID-19 diagnosis were 0.56 (95% CI 0.51-0.60) and 0.60 (95% CI 0.54-0.65), respectively. For CT scans these were 0.85 (95% CI 0.79-0.90) and 0.50 (95% CI 0.41-0.60), respectively. This gave a statistically significant mean increase in sensitivity with CT compared with CXR, of 29% (95% CI 19%-38%, p<0.0001). Specificity was not significantly different between the two modalities.

Chest X-ray findings were not statistically significantly or clinical meaningfully associated with vital signs, laboratory parameters or 30-day outcomes.

**Conclusions:** Computed tomography has substantially improved diagnostic performance over CXR in COVID-19. CT should be strongly considered in the initial assessment for suspected COVID-19. This gives potential for increased sensitivity and considerably faster turnaround time, where capacity allows and balanced against excess radiation exposure risk.

## Introduction

SARS-CoV 2 and its resulting disease, COVID-19, have propagated exponentially worldwide, with over 10 million cases in 188 countries at the time of writing [1,2].

The gold standard for diagnosis of the virus is the detection of viral RNA through reverse transcriptase polymerase chain reaction (RT-PCR) of respiratory tract samples. However, this method has several limitations including: (1) low sensitivity at 59-71% [3,4], (2) relatively slow turnaround times ranging from a few hours to several days [5], (3) high expense and (4) limited capacity for testing in many countries.

Computed tomography (CT) has been shown to be more sensitive than RT-PCR for diagnosis of COVID-19 [3,4], while being significantly faster and cheaper. This comes with a large radiation dose and capacity is still lacking in many countries.

Plain film chest X-ray (CXR) is ubiquitous worldwide, with a 30-70x lower dose of radiation[6] and is commonly performed as an initial investigation in COVID-19.

Studies have so far only evaluated imaging in those with confirmed infection, it is therefore, not possible to calculate the specificity of these modalities. In the context of the global pandemic, infection may be widespread in the community, often with subclinical infection [7,8]. A reliable and rapid method to detect infection in the general population, who may present to medical personnel with other complaints, is needed.

Despite its extensive use, the specificity and sensitivity of CXR in the general emergency population for diagnosis of COVID-19 is unknown, nor how imaging features correlate with severity.

This study evaluated the performance of CXR in diagnosing COVID-19 in the emergency department (ED) of a tertiary care hospital.

## Methods

This study was conducted at the Royal Free Hospital, London, UK, an academic health science centre and nationally designated centre for High Consequence Infectious Diseases [9].

All individuals attending the emergency department who had paired posterior-anterior chest radiographs and RT-PCR nasopharyngeal swabs for COVID-19 at the time of initial attendance between 16^th^ March 2020 and 16^th^ April 2020 were included.

All chest radiographs were reported by a Consultant Radiologist and rated on an ordinal scale for probability of COVID-19: Alternative pathology identified, not COVID-19; Clear chest, unlikely COVID; Indeterminate findings for COVID-19; Classical findings of COVID-19, based on the British Society of Thoracic Imaging’s (BSTI) reporting templates (table 1) [10]. These were reported prior to RT-PCR results being available.

**Table 1.**
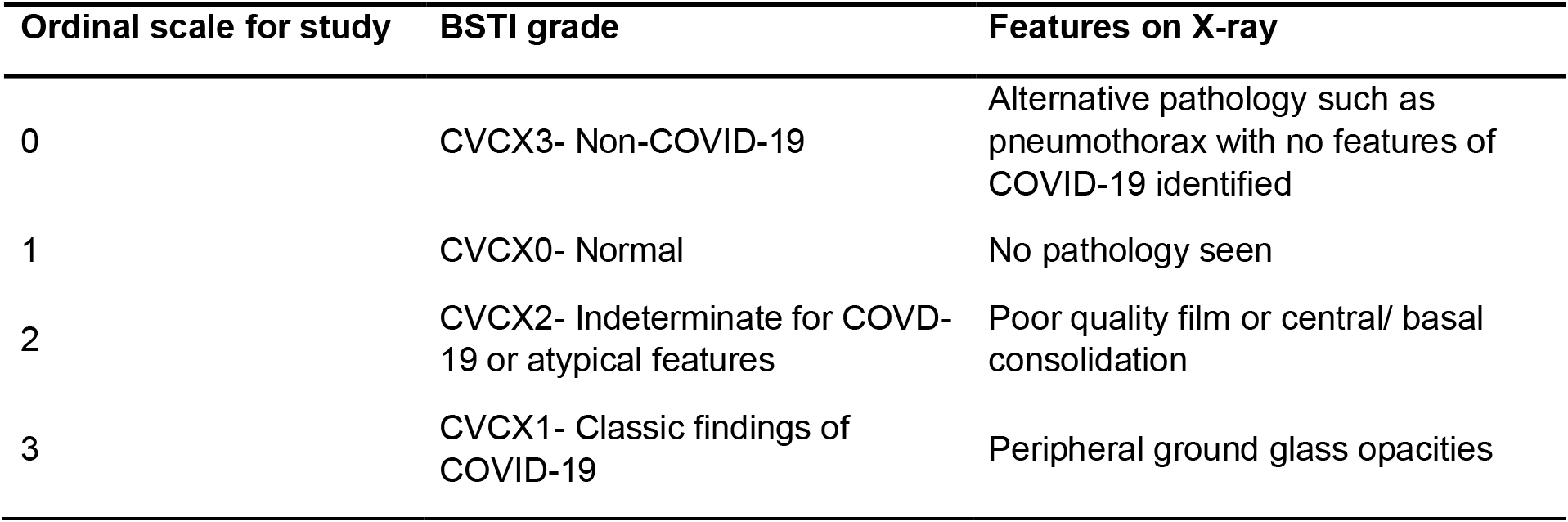
Ordinal scale used in this study based on the British Society of Thoracic Imaging (BSTI) Reporting Template [10]

RT-PCR of swabs were performed in laboratories either at our centre or at a public health laboratory (PHE Collindale, UK), according to published national standard operating procedures [11]. Subsequent RT-PCR swabs taken within 30 days of initial ED attendance were also included.

CT scans performed within 30 days of attendance were retrieved. These were also reported according to the BSTI template. CT pulmonary angiogram was performed in the ED if the D-dimer was >5000 to exclude pulmonary emboli as per the locally agreed protocol. Subsequent CT chest imaging (whether pulmonary angiogram, contrast or non-contrast) was performed on the basis of clinical suspicion.

Prospectively recorded data was extracted from the Cerner Millennium electronic patient record system (Cerner Corp., Kansas City, MO).

### Primary Outcome

The primary outcome is sensitivity and specificity of initial CXR, where it is reported as having classic COVID-19 features in the ED. This is compared with RT-PCR swab as the reference standard for diagnosis of COVID-19.

In the event of multiple RT-PCR swabs during one attendance, a single positive swab was taken as an overall positive test during one admission.

### Secondary Outcomes

In those patients who also had CT scans of the thorax, the diagnostic accuracy was compared with CXR, with RT-PCR again as the reference standard. Sensitivity and specificity of CXR when X-rays reported as indeterminate or atypical for COVID-19 were classed as positive was also calculated.

Chest x-ray findings were correlated with vital signs at attendance and blood results, including: neutrophil counts, D-dimer and C-reactive protein, which have been associated with poor prognosis in COVID-19 [12]. Hazard ratios for clinical outcomes including direct admission to the intensive treatment unit (ITU) from ED and 30-day mortality rates were also calculated for CXR reporting categories.

### Statistical Analysis

In the event of missing data, multiple imputation was conducted using a Predictive Mean Matching algorithm, via the MICE R package, as described previously [13]. Briefly, this uses a linear regression model (or logistic regression model for categoric data), to find a random value based on already observed data, to replace missing fields [14]. Variables without missing data fields were not modified. The number of imputed datasets was similar in number to the percentage of missing data as suggested by White and colleagues [15]. Balance diagnostics with density plots are available in supplementary file 1, adequate balance was assessed via visual inspection of imputed distributions with respect to the original dataset.

The propensity for a CXR being reported as positive or negative for COVID-19 was calculated for several plausible covariates that may influence image characteristics such as Age, Gender, Ethnicity, pre-existing morbidities and the respiratory rate of the patient using a generalised linear model [16]. X-ray positive and negative groups were then matched in each imputed dataset using the nearest neighbour algorithm, with a calliper of 0.2 of the propensity score standard deviation, without replacement and in random sequential order to obtain a 1:1 match as described elsewhere [17].

The balance of the match data was assessed quantitatively with mean differences of covariates in each of the X-ray groups pre- and post-matching, with a difference of less than 0.1% considered a good match (supplementary figure 2). Visual inspection of matches was also conducted to ensure balance (supplementary figures 2, 3 and 4).

After matching, outcome data were adjusted for covariates including age, gender, ethnicity and presence of co-morbidities as well as C-reactive protein, D-dimer, troponin and vital signs. This was achieved by generalised linear regression for continuous outcome data, binomial logistic regression for binary categoric outcomes, or ordinal logistic regression in the case of CXR where it is the outcome variable.

These regression models were run on each imputed dataset and outcomes were pooled together across each imputed data set according to Rubin’s rules [18] to give an overall estimate.

### Diagnostic Accuracy Statistics

Chest X-rays reported as classical for COVID-19 as per the BSTI guidelines were considered a positive test in the primary analysis. In a secondary analysis X-rays reported as ‘Indeterminate’ or ‘Atypical’ for COVID-19 were also considered positive. All other reports were classified as a negative test. These were compared to nasopharyngeal aspirate RT-PCR results, which were taken as the gold standard for diagnosis of COVID-19. Where more than one swab was taken during the study period (up to 30 days after initial attendance), a single positive result was taken as a positive result for calculation of diagnostic accuracy statistics.

Sensitivity, specificity, predictive values and diagnostic accuracy were calculated using the propensity matched data after imputation and pooled across imputed datasets with 95% confidence intervals. Apparent and true prevalence based on this dataset are also given for interpretation of the predictive values.

Chest CTs were also reported according to the BSTI guidelines as with X-ray. Diagnostic statistics were calculated on raw, unmatched and non-imputed data (due to a low volume of data for imputation and matching) in the same manner as X-ray. Mean differences and 95% confidence intervals between CT and X-ray for each of the diagnostic statistics are given, with a p-value calculated from the confidence intervals.

Agreement between the modalities was assessed on the unmatched dataset, in the sample where CT, CXR and RT-PCR were all available using Cohen’s (for two group agreement) and Fleiss’ Kappa (when all 3 are compared).

### Data Presentation

Descriptive statistics are given as means and standard deviations for normally distributed data and as medians and interquartile ranges for non-normally distributed data, before and after matching and multiple imputation (for the latter these statistics are pooled across imputations).

Association of explanatory variables with SARS-CoV 2 and Chest X-ray findings are given as odds ratios in uni- and multi-variate configurations.

Data was considered statistically significant if p < 0.05. Given the large number of analyses in this paper, data is separately highlighted if p<0.001 as a secondary threshold to address the potential for false positives with multiple testing.

Analyses were conducted using R 4.0.0 (R Foundation for Statistical Computing, Vienna, Austria) and code for the analyses is given in supplementary file 2.

### Sample size calculation

In this study, the lower confidence interval for sensitivity of CXR as reported by Wong et al.[19] (56%) was used as an estimate of likely sensitivity for COVID-19. A power of 80% at an alpha of 0.05 was used to calculate the sample size for sensitivities and specificities of 56%. This gave an estimated sample size of 165 in each of the COVID-19 negative and positive groups by RT-PCR (total 330).

### Ethical approval

This study was registered with the local institutional review board as a service evaluation using anonymised data only. No formal ethics committee review was required.

### Reporting Guidelines

This study is reported according to the STARD guidelines [20] for diagnostic accuracy studies.

## Results

1,198 eligible patients with both CXR and RT-PCR were identified in the study period (figure 1). Their characteristics, stratified by positivity for SARS-CoV 2 infection by RT-PCR is summarized in table 2. This showed that those with confirmed SARS-CoV 2 infection were more likely to be male, older (mean age 66.2 vs 62.7), have lower saturations, higher respiratory rates, whilst being more likely to be admitted and die within 30 days. There was a signification association with X-ray images and SARS-CoV 2 at baseline, with 59.6% having classic imaging features of COVID-19 in those with positive swabs versus 39.1% in those with negative swabs. There was 8.6% missing data overall in the dataset when variables with >50% missing data were removed and 15 imputations were performed on these remaining variables only.

**Table 2.**
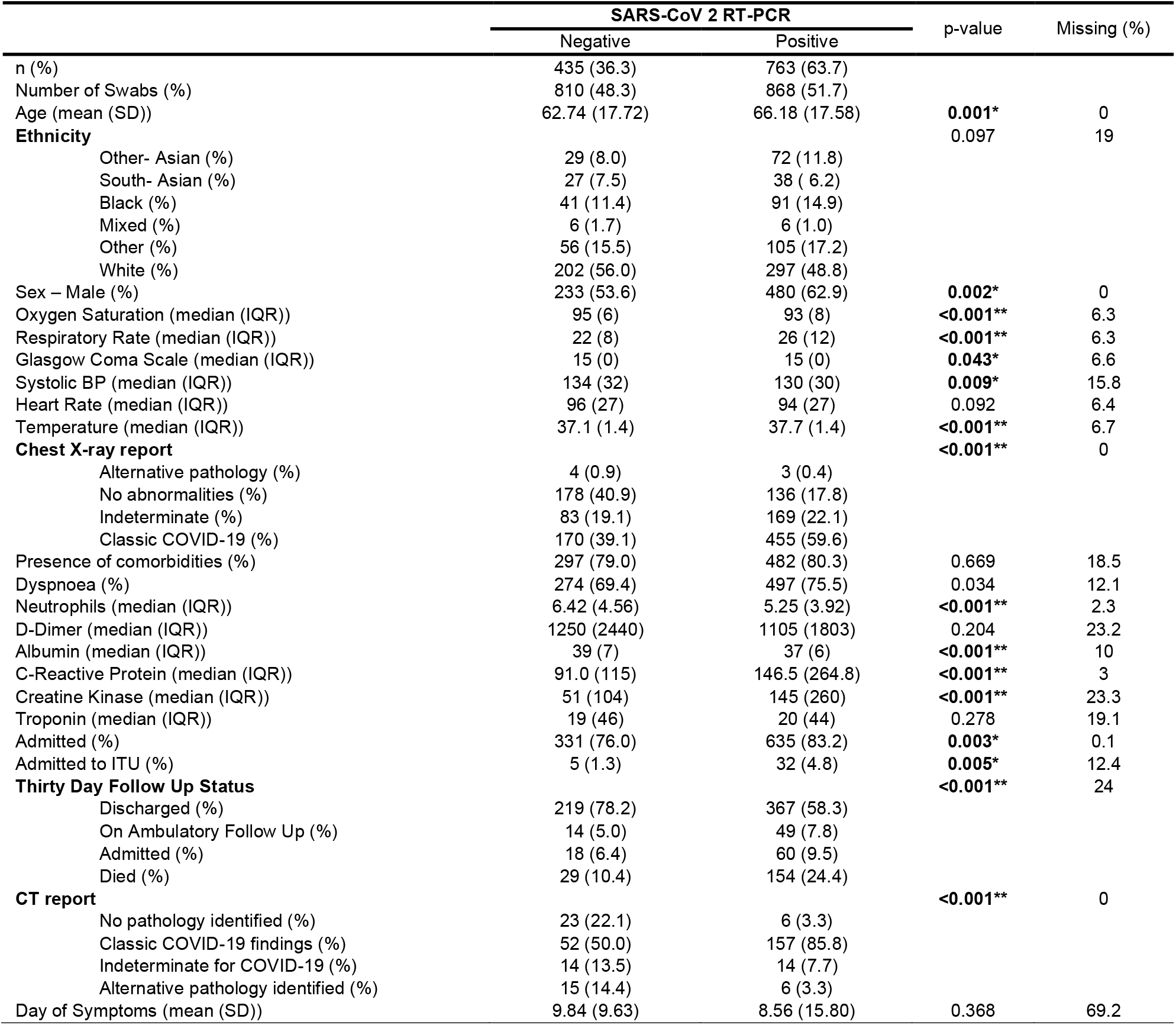
Baseline characteristics of dataset stratified by overall SARS-CoV 2 RT-PCR status, including subsequent swabs during the study period-NB there were 480 additional swabs on 399 unique patients with a median of 2 and mean of 3.5 per patient; *significant at p< 0.05; **significant at p< 0.001

**Table 3.**
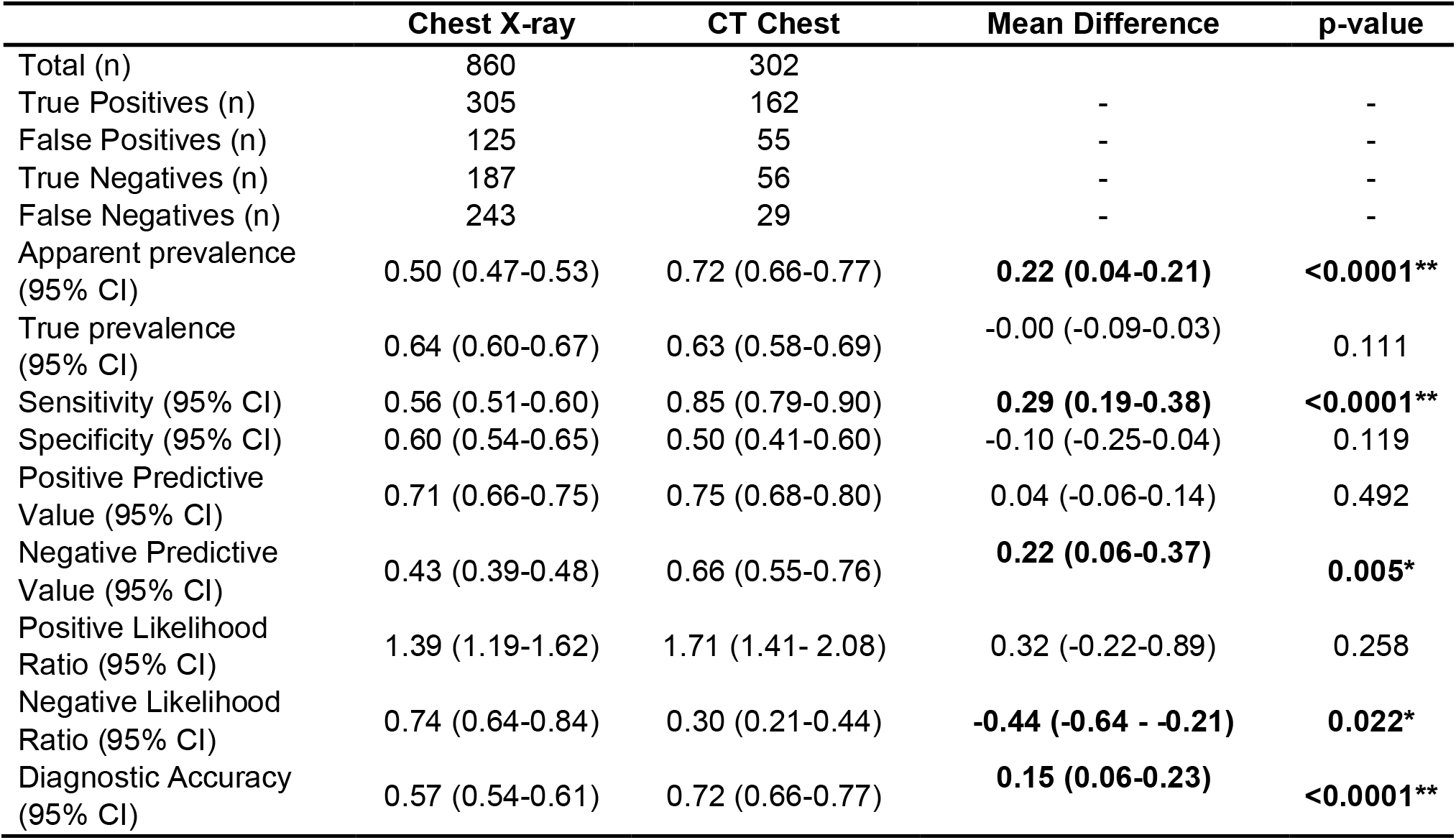
Diagnostic Accuracy Metrics for CXR and CT Chest with RT-PCR for SARS-CoV 2, as the reference standard; *significant difference at the <0.05 level; **significant difference at the <0.0001 level

|  | Chest X-ray | CT Chest | Mean Difference | p-value |
| --- | --- | --- | --- | --- |
| Total (n) | 860 | 302 |  |  |
| True Positives (n) | 305 | 162 | - | - |
| False Positives (n) | 125 | 55 | - | - |
| True Negatives (n) | 187 | 56 | - | - |
| False Negatives (n) | 243 | 29 | - | - |
| Apparent prevalence (95% CI) | 0.50 (0.47-0.53) | 0.72 (0.66-0.77) | **0.22 (0.04-0.21)** | **<0.0001\*\*** |
| True prevalence (95% CI) | 0.64 (0.60-0.67) | 0.63 (0.58-0.69) | -0.00 (-0.09-0.03) | 0.111 |
| Sensitivity (95% CI) | 0.56 (0.51-0.60) | 0.85 (0.79-0.90) | **0.29 (0.19-0.38)** | **<0.0001\*\*** |
| Specificity (95% CI) | 0.60 (0.54-0.65) | 0.50 (0.41-0.60) | -0.10 (-0.25-0.04) | 0.119 |
| Positive Predictive Value (95% CI) | 0.71 (0.66-0.75) | 0.75 (0.68-0.80) | 0.04 (-0.06-0.14) | 0.492 |
| Negative Predictive Value (95% CI) | 0.43 (0.39-0.48) | 0.66 (0.55-0.76) | **0.22 (0.06-0.37)** | **0.005\*** |
| Positive Likelihood Ratio (95% CI) | 1.39 (1.19-1.62) | 1.71 (1.41- 2.08) | 0.32 (-0.22-0.89) | 0.258 |
| Negative Likelihood Ratio (95% CI) | 0.74 (0.64-0.84) | 0.30 (0.21-0.44) | **-0.44 (-0.64 - -0.21)** | **0.022\*** |
| Diagnostic Accuracy (95% CI) | 0.57 (0.54-0.61) | 0.72 (0.66-0.77) | **0.15 (0.06-0.23)** | **<0.0001\*\*** |

**Figure 1.**
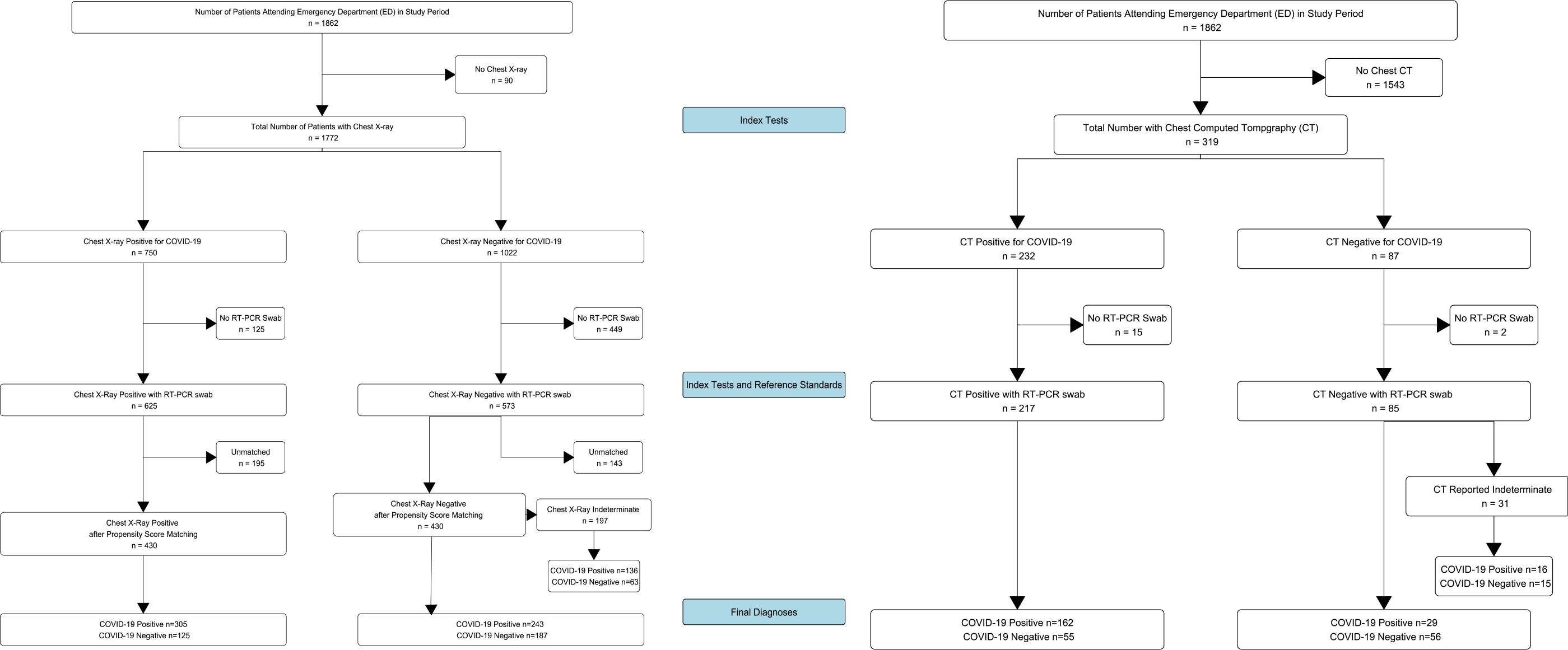
Inclusion and exclusion of patients during study period with test results

After multiple imputation for missing data and pooled propensity score matching for plausible covariates that may affect CXR reporting, there were 430 patients in each of the X-ray positive and X-ray negative groups, for a total of 860 patients. Adequate balance was achieved for relevant covariates with a mean difference of <0.1 between groups (supplementary table 2).

Computed tomography (CT) was performed in 302 patients with paired RT-PCR during the same time period, with a median serial interval of 4.5 days (inter quartile range 0-17) after the initial attendance in ED and of these 30.1% were within one day of attendance.

### Diagnostic Accuracy

The pooled sensitivity and specificity of CXR was 0.56 (95% CI 0.51-0.60) and 0.60 (95% CI 0.54-0.65), respectively (table 4). This gave an overall diagnostic accuracy of 0.57 (95% CI 0.54-0.61) for CXR.

**Table 4.** Association of covariates with RT-PCR status for SARS-CoV 2, following propensity score matching and binomial logistic regression; SD-Standard deviation; IQR-Interquartile Range; *p<0.05; **p<0.001

|  |  | SARS-CoV 2 RT-PCR |  | OR (univariable) | OR (multivariable) |
| --- | --- | --- | --- | --- | --- |
|  |  | Negative | Positive |  |  |
| n |  | 312 | 548 |  |  |
| Chest X-ray report | Alternative pathology (%) | 3 (0.8) | 3 (0.5) | - | - |
|  | No abnormalities (%) | 123 (39.6) | 104 (19.1) | 0.76 (0.08-6.82, p=0.801) | 0.48 (0.03-8.82, p=0.620) |
|  | Indeterminate/ atypical findings (%) | 61 (19.5) | 136 (4.8) | 1.99 (0.22-17.81, p=0.535) | 0.92 (0.05-16.88, p=0.952) |
|  | Classic COVID (%) | 125 (40.1) | 305 (55.6) | 2.17 (0.24-19.19, p=0.484) | 1.14 (0.06-20.98, p=0.927) |
| Age | Mean (SD) | 61.8 (17.9) | 67.0 (17.7) | **1.02 (1.01-1.02, p<0.001)\*\*** | **1.02 (1.00-1.03, p=0.028)\*** |
| Sex | Female (%) | 138 (44.3) | 212 (38.7) | - | - |
|  | Male (%) | 174 (55.7) | 336 (61.3) | 1.26 (0.93-1.70, p=0.137) | 1.19 (0.83-1.71, p=0.340) |
| Ethnicity | Other Asian (%) | 31 (9.9) | 66 (12.0) | - | - |
|  | White (%) | 164 (52.7) | 270 (49.2) | 0.76 (0.44-1.31, p=0.326) | 0.73 (0.38-1.40, p=0.339) |
|  | Black (%) | 39 (12.4) | 84 (15.3) | 1.01 (0.52-1.98, p=0.974) | 0.92 (0.43-1.97, p=0.827) |
|  | Mixed (%) | 6 (1.8) | 4 (0.8) | 0.36 (0.08-1.62, p=0.184) | 0.74 (0.11-4.94, p=0.754) |
|  | South Asian (%) | 22 (7.0) | 36 (6.6) | 0.77 (0.34-1.76, p=0.531) | 0.68 (0.28-1.65, p=0.390) |
|  | Other (%) | 51 (16.2) | 89 (16.2) | 0.82 (0.43-1.55, p=0.535) | 0.88 (0.45-1.74, p=0.716) |
| Comorbidity | No (%) | 65 (20.8) | 95 (17.4) | - | - |
|  | Yes (%) | 247 (79.2) | 453 (82.6) | 1.25 (0.82-1.89, p=0.296) | 1.00 (0.53-1.88, p=0.993) |
| Dyspnoea on attendance | No (%) | 90 (28.8) | 139 (25.4) | - | - |
|  | Yes (%) | 222 (71.2) | 409 (74.6) | 1.19 (0.82-1.73, p=0.356) | 0.84 (0.53-1.32, p=0.447) |
| Oxygen Saturation | Median (IQR) | 96 (6) | 93 (8) | **0.94 (0.91-0.97, p<0.001)\*\*** | 0.97 (0.93-1.00, p=0.072) |
| Respiratory rate | Median (IQR) | 23 (8) | 25 (8) | **1.04 (1.01-1.07, p=0.002)\*** | 1.01 (0.98-1.05, p=0.462) |
| Glasgow Coma Scale | Median (IQR) | 15 (0) | 15 (0) | 1.02 (0.89-1.17, p=0.819) | 1.21 (0.98-1.48, p=0.073) |
| Temperature | Mean (SD) | 37.2 (1.4) | 37.7 (1.1) | **1.48 (1.26-1.73, p<0.001)\*\*** | **1.44 (1.20-1.74, p<0.001)\*\*** |
| Heart Rate | Mean (SD) | 96.7 (20.5) | 94.9 (21.5) | 1.00 (0.99-1.00, p=0.305) | 1.00 (0.99-1.01, p=0.702) |
| Systolic Blood Pressure | Mean (SD) | 136.2 (25.8) | 132.6 (24.5) | 0.99 (0.99-1.00, p=0.086) | 0.99 (0.98-1.00, p=0.097) |
| Neutrophils | Median (IQR) | 6.26 (4.52) | 5.05 (3.93) | **0.92 (0.89-0.96, p<0.001)\*\*** | **0.87 (0.82-0.91, p<0.001)\*\*** |
| D-Dimer | Median (IQR) | 1220 (2343) | 1061 (1814) | 1.00 (1.00-1.00, p=0.403) | 1.00 (1.00-1.00, p=0.419) |
| C-Reactive Protein | Median (IQR) | 45 (100) | 77 (107) | **1.00 (1.00-1.01, p<0.001)\*\*** | **1.00 (1.00-1.01, p=0.021)\*** |
| Troponin | Median (IQR) | 20 (55) | 21 (46) | 1.00 (1.00-1.00, p=0.890) | 1.00 (1.00-1.00, p=0.667) |
| Albumin | Median (IQR) | 39 (7) | 37 (6) | 0.97 (0.94-1.00, p=0.071) | 1.02 (0.98-1.06, p=0.432) |
| Creatine Kinase | Median (IQR) | 94 (131) | 145 (263) | 1.00 (1.00-1.00, p=0.119) | 1.00 (1.00-1.00, p=0.152) |
| Admitted from ED | Admitted (%) | 235 (75.2) | 453 (82.7) | - | - |
|  | Discharged (%) | 77 (24.8) | 95 (17.3) | **1.56 (1.06-2.33, p=0.022)\*\*** | 1.35 (0.79-2.30, p=0.272) |
| Admitted To ITU from ED | No (%) | 307 (98.5) | 532 (97.1) | - | - |
|  | Yes (%) | 5 (1.5) | 16 (2.9) | 1.92 (0.60-6.18, p=0.274) | 1.06 (0.25-4.40, p=0.940) |
| Thirty Day Follow up Status | Discharged (%) | 259 (83.0) | 368 (67.1) | - | - |
|  | Admitted (%) | 22 (6.9) | 47 (8.5) | 1.53 (0.82-2.87, p=0.181) | 1.64 (0.77-3.51, p=0.198) |
|  | Dead (%) | 31 (10.1) | 133 (24.4) | **3.00 (1.86-4.84, p<0.001)\*\*** | **2.81 (1.22-6.50, p=0.017)\*** |

In comparison, sensitivity and specificity for CT was 0.85 (95% CI 0.79-0.90) and 0.50 (95% CI 0.41-0.60), respectively. This gave a statistically significant mean increase in sensitivity with CT compared with CXR by 29% (95% CI 19%-38%, p<0.0001). Specificity was not significantly different between the two modalities. Diagnostic accuracy and negative predictive values were also significantly increased with CT at 0.15 and 0.22, respectively, while the negative likelihood ratio was significantly decreased at −0.44. This shows that the post-test odds of being negative for SARS-CoV 2 by RT-PCR with a negative CT is significantly lower.

Taking X-rays reported as indeterminate as positive increased the sensitivity of CXR to 0.80 (95% CI 0.77-0.84), however reduced specificity to 0.40 (95% CI 0.35-0.46). When CT scans reported as indeterminate are also considered positive the sensitivity of CT increased to 0.93 (95% CI 0.89-0.96), whilst mean specificity reduced to 0.37 (95% CI 0.28-0.47), although this was not statistically different from when indeterminate CTs are considered negative. Sensitivity of CT remained significantly higher than CXR (when indeterminates are considered positive for both) by 0.13 (95% CI 0.05-0.19, p<0.001), specificity was not significantly different between the two.

When comparing only the unimputed, unmatched subset of data where CT, RT-PCR and CXR were all performed (n=287), the agreement between CT and CXR was poor (Cohen’s kappa 0.406). Agreement between all three modalities was also poor (Fleiss’ kappa 0.361).

### Association of CXR with Markers of Severity and Outcomes

Association of covariates with RT-PCR results is shown in table 4 and figure 2. Those who tested positive for SARS-CoV 2 by RT-PCR were significantly more likely to have a classical X-ray (OR 1.79 95% CI 1.25-2.56, p<0.002) as would be expected by the diagnostic accuracy statistics (table 4). When the CXR report is considered as an ordered scale, worsening grades of report were associated more strongly with RT-PCR positivity, with a 1.94 × increase in odds for each grade.

**Figure 2.**
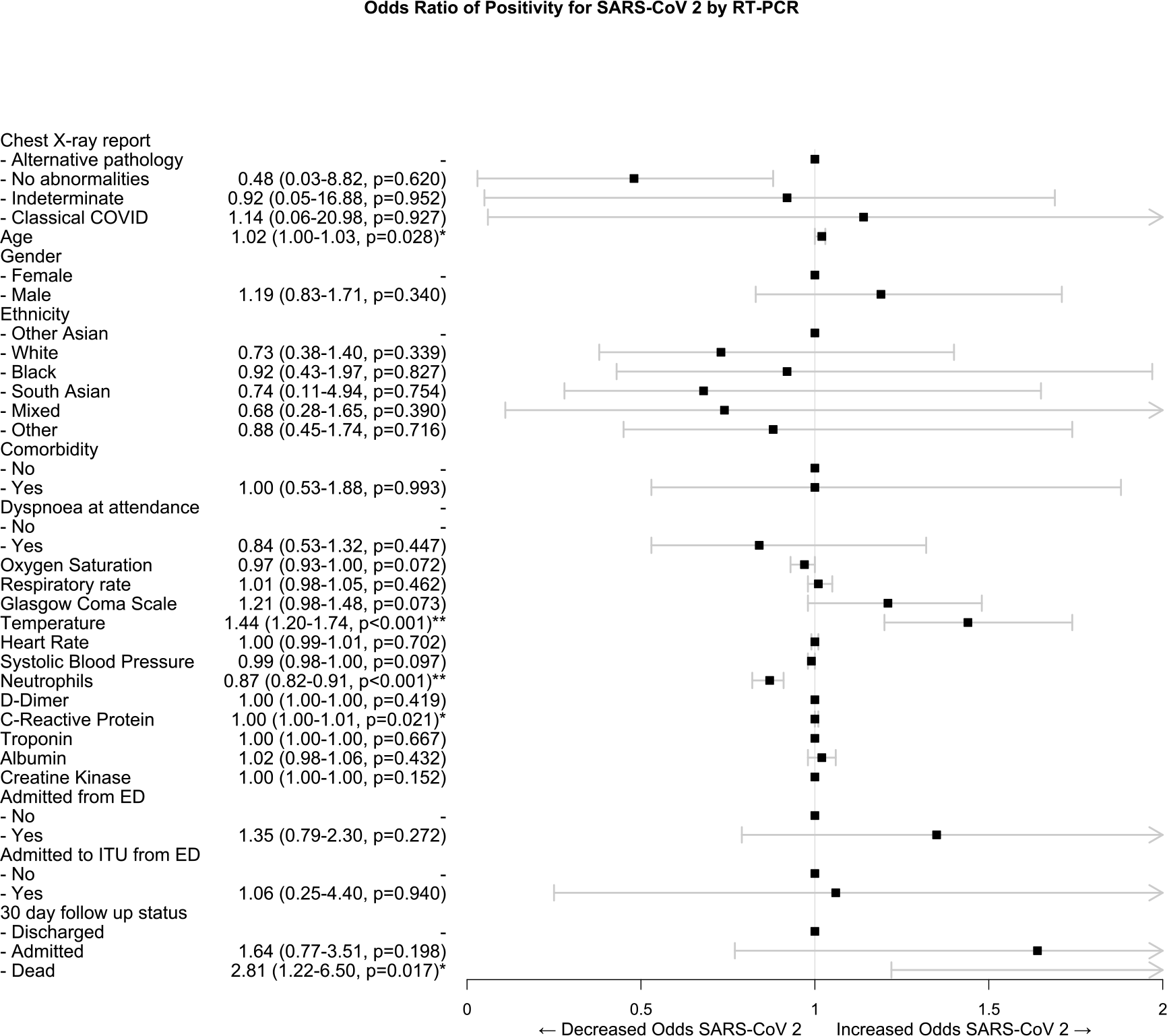
Forest plot of odds ratios of variables associated with RT-PCR positivity for SARS-CoV 2, following multiple imputation, propensity score matching and binomial logistic regression; *significant difference at the <0.05 level; **significant difference at the <0.001 level

Positive chest X-rays for COVID-19 were significantly associated with lower oxygen saturations (OR 0.94 95% CI 0.92-0.97, p<0.001) and temperatures (2.30 95% CI 1.46-3.63, p<0.001) in the ED following propensity score matching and multivariate regression (table 5 and figure 3).

**Table 5.** Association of covariates with CXR report following propensity score matching and either binomial or ordinal logistic regression; SD-Standard deviation; IQR-Interquartile Range; *p<0.05; **p<0.001

|  |  | X-ray report |  | OR (univariable) | OR with XR as binary outcome (multivariable) | OR with XR as ordinal variable (multivariable) |
| --- | --- | --- | --- | --- | --- | --- |
|  |  | Other X-ray Findings | Classical COVID-19 |  |  |  |
| n |  | 430 | 430 |  |  |  |
| RT-PCR for SARS-CoV 2 | Negative (%) | 187 (43.4) | 125 (29.1) | - | - | - |
|  | Positive (%) | 243 (56.6) | 305 (70.9) | **1.85 (1.36-2.56, p<0.001)\*\*** | **1.79 (1.25-2.56, p<0.002)\*** | **1.94 (1.37-2.76, p<0.001)\*\*** |
| Age | Mean (SD) | 65.0 (18.9) | 65.3 (16.9) | 1.00 (0.99-1.01, p=0.849) | 0.99 (0.98-1.00, p=0.164) | 1.00 (0.99-1.01, p=0.542) |
| Sex | Female (%) | 176 (40.9) | 175 (40.6) | - | - | - |
|  | Male (%) | 254 (59.1) | 255 (59.3) | 1.01 (0.75-1.37, p=0.940) | 0.87 (0.63-1.20, p=0.400) | 1.02 (0.49-2.09, p=0.967) |
| Ethnicity | Other Asian (%) | 49 (11.4) | 48 (11.2) | - | - | - |
|  | South Asian (%) | 29 (6.7) | 29 (6.7) | 1.04 (0.52-2.04, p=0.912) | 1.02 (0.47-2.17, p=0.965) | 1.02 (0.49-2.09, p=0.967) |
|  | Black (%) | 61 (14.2) | 61 (14.2) | 1.02 (0.55-1.85, p=0.957) | 0.88 (0.46-1.69, p=0.719) | 0.92 (0.52-1.65, p=0.789) |
|  | Mixed (%) | 5 (1.2) | 5 (1.2) | 0.92 (0.21-4.00, p=0.911) | 0.86 (0.18-4.17, p=0.853) | 0.85 (0.17-4.30, p=0.838) |
|  | Other (%) | 70 (16.3) | 70 (16.3) | 1.02 (0.58-1.79, p=0.943) | 0.98 (0.52-1.82, p=0.942) | 0.93 (0.53-1.64, p=0.810) |
|  | White (%) | 216 (50.2) | 217 (50.5) | 1.03 (0.63-1.67, p=0.913) | 0.97 (0.57-1.67, p=0.926) | 0.90 (0.55-1.47, p=0.666) |
| Comorbidity | No (%) | 82 (19.1) | 78 (18.1) | - | - | - |
|  | Yes (%) | 348 (80.9) | 352 (81.9) | 0.95 (0.66-1.36, p=0.777) | 0.93 (0.59-1.49, p=0.782) | 0.88 (0.57-1.37, p=0.592) |
| Dyspnoea | No (%) | 191 (29.3) | 103 (24.0) | - | - | - |
|  | Yes (%) | 304 (70.7) | 327 (76.0) | 1.31 (0.92-1.88, p=0.123) | 1.20 (0.80-1.82, p=0.380) | 1.22 (0.83-1.80, p=0.301) |
| Oxygen Saturation | Median (IQR) | 95 (7) | 93 (7) | **0.94 (0.91-0.96, p<0.001)\*\*** | **0.94 (0.92-0.97, p<0.001)\*\*** | **0.94 (0.91-0.97, p<0.001)\*\*** |
| Respiratory rate | Median (IQR) | 24 (10) | 24 (10) | 1.01 (0.99-1.02, p=0.570) | 0.97 (0.94-1.00, p=0.063) | 0.98 (0.96-1.01, p=0.157) |
| Glasgow Coma Scale | Median (IQR) | 15 (0) | 15 (0) | 1.04 (0.92-1.19, p=0.524) | 1.05 (0.90-1.23, p=0.503) | 1.05 (0.92-1.21, p=0.464) |
| Temperature | Mean (SD) | 37.6 (1.1) | 37.5 (1.3) | 0.93 (0.83-1.06, p=0.297) | **0.79 (0.67-0.93, p=0.006)\*** | **0.85 (0.73-0.99, p=0.031)\*** |
| Heart Rate | Mean (SD) | 95.7 (21.4) | 95.5 (21.0) | 1.00 (0.99-1.01, p=0.888) | 1.00 (0.99-1.01, p=0.864) | 1.00 (0.99-1.01, p=0.872) |
| Systolic Blood Pressure | Mean (SD) | 133.8 (25.0) | 134.0 (25.6) | 1.00 (0.99-1.01, p=0.907) | 1.00 (0.99-1.01, p=0.335) | 1.00 (1.00-1.01, p=0.478) |
| Neutrophils | Median (IQR) | 5.44 (4.54) | 5.67 (4.03) | 1.00 (0.97-1.04, p=0.892) | 0.96 (0.92-1.01, p=0.143) | 0.96 (0.92-1.01, p=0.115) |
| D-Dimer | Median (IQR) | 1119 (2221) | 1119 (1850) | 1.00 (1.00-1.00, p=0.513) | 1.00 (1.00-1.00, p=0.568) | 1.00 (1.00-1.00, p=0.385) |
| C-Reactive Protein | Median (IQR) | 46 (93) | 88 (110) | **1.00 (0.99-1.00, p<0.001)\*\*** | **1.00 (1.00-1.01, p<0.001)\*\*** | **1.00 (1.00-1.01, p<0.001)\*\*** |
| Troponin | Median (IQR) | 23 (54) | 20 (46) | 1.00 (1.00-1.00, p=0.231) | 1.00 (1.00-1.00, p=0.277) | 1.00 (1.00-1.00, p=0.059) |
| Albumin | Median (IQR) | 39 (7) | 37 (6) | **0.93 (0.90-0.96, p<0.001)\*\*** | **0.93 (0.90-0.97, p=0.001)\*** | **0.94 (0.91-0.97, p=0.001)\*** |
| Creatine Kinase | Median (IQR) | 110 (183) | 134 (239) | 1.00 (1.00-1.00, p=0.535) | 1.00 (1.00-1.00, p=0.242) | 1.00 (1.00-1.00, p=0.186) |
| Admitted from ED | Admitted (%) | 315 (73.3) | 373 (86.7) | **2.37 (1.63-3.46, p<0.001)\*\*** | **2.30 (1.46-3.63, p<0.001)\*\*** | **2.22 (1.47-3.33, p<0.001)\*\*** |
|  | Discharged (%) | 115 (26.7) | 57 (13.3) | - | - | - |
| Admitted to ITU from ED | No (%) | 423 (98.4) | 416 (96.7) | - | - | - |
|  | Yes (%) | 7 (1.6) | 14 (3.3) | 2.17 (0.69-6.67, p=0.181) | 1.27 (0.32-5.00, p=0.732) | 1.34 (0.36-5.00, p=0.653) |
| 30 Day Follow Up Status | Discharged (%) | 316 (73.5) | 311 (72.3) | - | - | - |
|  | Admitted (%) | 34 (7.9) | 34 (7.9) | 1.31 (0.81-2.13, p=0.282) | 1.32 (0.69-2.53, p=0.392) | 1.43 (0.78-2.63, p=0.653) |
|  | Dead (%) | 80 (18.6) | 85 (19.8) | 1.03 (0.73-1.45, p=0.886) | 1.38 (0.80-2.37, p=0.247) | 1.41 (0.87-2.27, p=0.157) |

**Figure 3.**
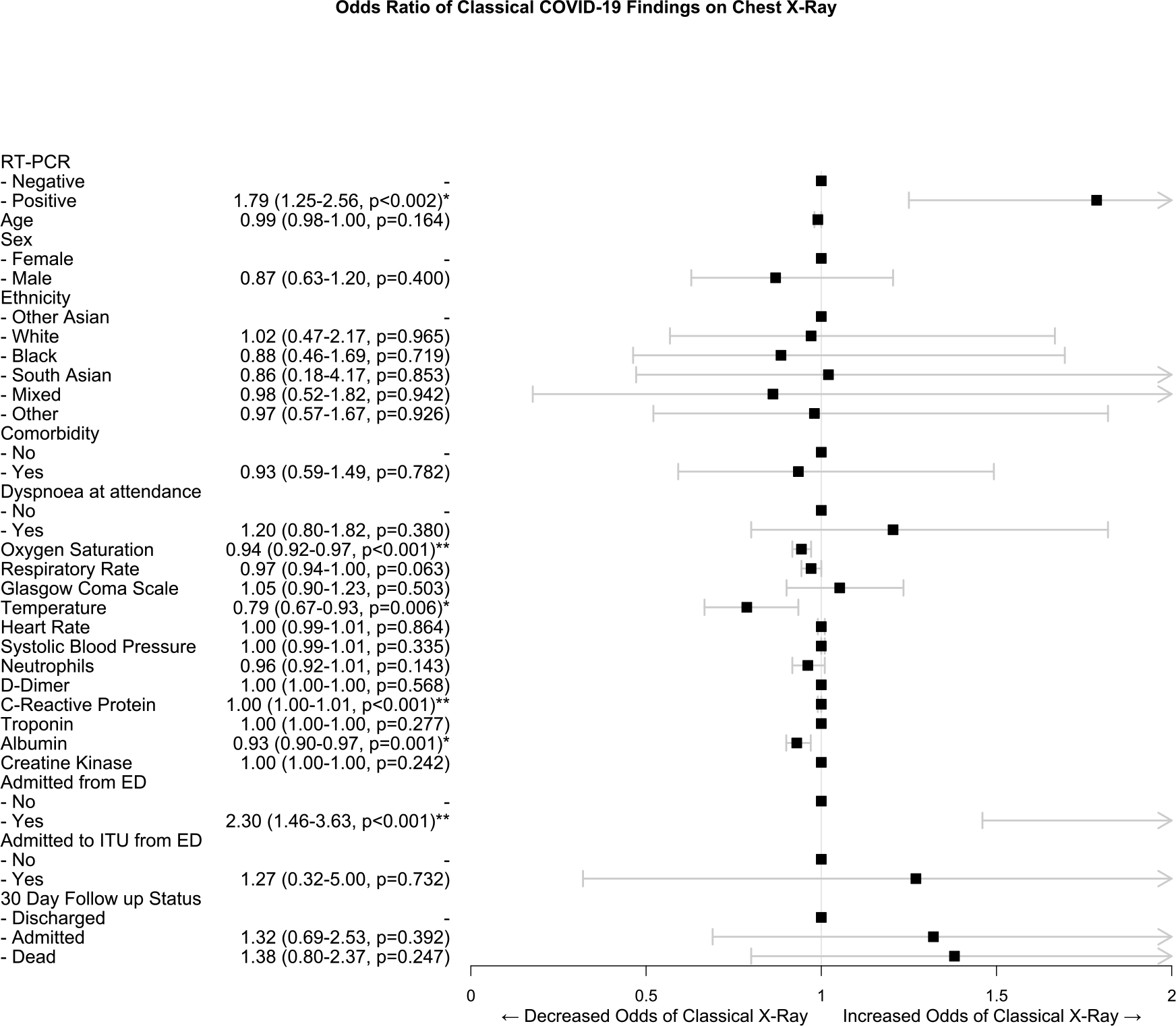
Forest plot of odds ratios of variables associated with classical Chest X-ray features COVID-19 following propensity score matching and binomial logistic regression; *significant difference at the <0.05 level; **significant difference at the <0.001 level

They also had higher rates of admission to a general ward from the ED (OR 2.30 95% CI 1.46-3.63, p<0.001) but no significant association with 30 day outcomes. There was a statistically significant increase in C-reactive protein with a positive X-ray, however, this is unlikely to be clinically meaningful due to the minimal association (OR 1.00 95% CI 1.00-1.01).

## Discussion

This study is the first to report the diagnostic accuracy of CXR and CT in the general emergency population during the COVID-19 pandemic.

We show that CXR has poor sensitivity and specificity for diagnosis of COVID-19, whilst CT has 29% higher sensitivity. Many international radiological guidelines advise against CT scanning for the initial assessment of COVID-19 [21–23] or where there are equivocal CXRs, whilst in other countries CT scanning is performed as a routine first line investigation. Our results suggest that CT should be considered in the initial assessment of COVID-19 and that CXR findings poorly correlate with CT findings in this setting. We also show that indeterminate and non-classical features of COVID-19 significantly increase the sensitivity of these imaging modalities, without a significant decrease in specificity. Further, we demonstrate the limited prognostic value of CXR in COVID-19.

These findings mirror what has previously been reported in the literature on individuals with confirmed COVID-19. Wong et al. [19] showed a sensitivity of 59% for initial X-ray in confirmed COVID-19 infection, similarly initial case series in China also reported a sensitivity of 59.1%[12].

A recent in press article from Italy reported a much higher sensitivity of 89% for CXR in a smaller general emergency population (n=535) without confirmed COVID-19 at attendance [24]. However, this used telephone follow up for clinical symptoms of COVID-19 as a reference standard in individuals with an initial negative RT-PCR swab and appeared to classify any abnormal X-ray as positive, which may inflate this figure. When indeterminate CXRs are counted as positive in this study, the sensitivity would be in line with this Italian data. In the US, a study of patients attending an urgent care centre with confirmed COVID-19, showed a much lower sensitivity at 41.7% for CXR where any abnormality was found on the images [25]. In this study 97/636 reports were re-classified from ‘possible pneumonia’ to ‘normal’ on second reading from a radiologist, highlighting the importance of inter-rater agreement and possibly explaining this low estimate.

Computed tomography has been reported in previous studies as being up to 98% sensitive for the diagnosis of COVID-19 in confirmed patients, when RT-PCR is used as the reference standard in confirmed patients [3,4]. These studies used any potential features of COVID-19 (e.g. ground glass opacification, crazy paving) as a positive scan, regardless of spatial distribution or features more characteristic of alternate pathology, unlike the BSTI guidelines used in this study. When we classified indeterminate CTs as positive like these latter studies, our estimates match their sensitivity values.

Consequently, a much lower specificity of 25% was found with initial RT-PCR in previous literature; however, it is reported that 10 out of 15 (67%) of these negatives subsequently tested positive. This would give an adjusted specificity of 75%, considering subsequent swabs as a reference standard, which combined with the wider CIs in these smaller studies, would bring estimates in line with the specificity in this paper. More recent meta-analyses have placed the pooled sensitivity of CT in populations with confirmed COVID-19 only, at 89.76% (95% CI 84.42%-93.84%) [26], in line with the estimates identified here.

There is limited coverage in the literature on association of X-ray findings with clinical and laboratory parameters and outcomes in the COVID-19 pandemic. This study demonstrates that classic appearances of COVID-19 were associated with initial lower saturations and lower temperature. Volume opacification of the lung fields were not quantified as a surrogate of severity; however, the use of the BSTI grading templates does this somewhat. When the X-ray report is considered as a graded scale from low likelihood of COVID-19 and severity to high likelihood and severity of disease there was no significant difference in association with vital signs or laboratory parameters compared with when the X-ray report is merely considered as a binary positive and negative outcome for COVID-19.

Borghesi and colleagues have devised a X-ray grading system, the Brixia score, for severity in admitted patients with confirmed SARS-CoV 2 infection [27]. They further found a significant increase in the severity of CXR by this scoring system in those who were discharged versus those who died [28,29].

Here, there were no relevant associations between CXR and laboratory values. This analysis also found no association with positive X-rays and 30 day outcomes after multivariate analyses, unlike Borghese et al. This is also in contrast to Guan et al. who found higher rates of ITU admission and death in those with positive imaging findings. However, these studies analysed only those with confirmed SARS-CoV 2 infection. The divergence observed in this study may be due to classifying those with ‘Alternate pathology/ Indeterminate’ or ‘CVXC3/ CVXC2’ as per the BSTI templates, negative for COVID-19 in these analyses. Other studies classified X-rays with any abnormality as a positive for COVID-19. These alternate distributions may still be reflective of underlying COVID-19 and we show significantly higher sensitivity for both CT and CXR when these are classed as positive. It may be that correlating indeterminate X-rays (in addition to classical images) with vitals, laboratory markers and 30 day outcomes would yield significant associations. However this may be unlikely, Xu and Zhang et al. found that those with classical bilateral and diffuse involvement in upper and lower lobes had more severe disease than those without [30,31].

There were a total of 70 confirmed pulmonary emboli (PEs) in our dataset out of 114 CT pulmonary angiograms (61.0%, 5.84% of all patients attending) performed in the emergency department. The incidence of venous thromboembolism is reported as ranging from 20-30% in admitted confirmed SARS-CoV 2 positive patients [32]. Although we have not focused on this cohort of patients in this paper for the sake of brevity and simplicity, this high incidence represents a further advantage for CT over CXR.

CT, even with the absence of contrast has been shown to have strong accuracy in the diagnosis of pulmonary emboli and many imaging features correlate with the presence of pulmonary emboli. Sensitivities of non-contrast CT for diagnosis of PE have been reported at 96.9% and specificity at 71.9% [33,34].

We therefore see the advantages of CT scanning in COVID-19 as threefold over other diagnostic techniques: 1) The rapid turnaround; 2) Increased sensitivity and 3) The possibility to identify pulmonary emboli in COVID-19, which are a significant burden in this group.

This must be balanced against the excess radiation exposure with CT. Radiation from CT and its association with carcinogenesis is difficult to quantify and no definitive epidemiological studies have confirmed excess risk of cancer[35]. Modern CT scanners and software reconstruction techniques continue to minimise radiation exposure and many ways of shielding parts of the body from radiation also exist. Nevertheless, the excess risk of lifetime cancer is estimated at 1 per 5,000 CT examinations[36].

### Strengths and Limitations

This study is the largest conducted on imaging in the COVID-19 pandemic and one of the only studies conducted in the general population during the pandemic rather than only in confirmed patients. This enables greater applicability to the clinical setting where the diagnosis is uncertain, in addition to being able to calculate specificity, which is not possible in most studies. This study was planned to be powered to detect a sensitivity and specificity of 56% for CXR and greatly exceeded the sample size necessary for this.

Comprehensive statistical analyses were conducted to account for confounders in both factors influencing reporting of CXR and in factors affecting outcomes. The data was collected from prospectively maintained electronic records; however, the retrieval took place retrospectively with its inherent disadvantages. We were not able to collect data on several relevant covariates such as specific comorbidities or markers of severity such as lymphocytes. Furthermore, there was a significant amount of missing data that required multiple imputation to replace, although the fit of this imputed data was good, actual, observed data would be ideal.

Inter-rater reliability of imaging reports was not analysed in this paper and there was the potential for individual radiologists to have greater or lesser accuracy in the diagnosis of COVID-19. The literature has so far suggested a strong degree of agreement between radiologists in reporting of COVID-19 images [28].

The single centre nature of this study further limits generalisability and the potential for inter-hospital disagreement in imaging, in addition to inter-rater disagreement.

Finally, the median time for patients to receive a CT scan was 4.5 days following initial attendance to ED. Thus, the scans may not have been directly comparable to the initial CXR, both because of the progression of disease and because the SARS-CoV 2 status may have been confirmed at this point, biasing the reporting of these scans.

### Future Research

Although this study used RT-PCR of nasopharyngeal swabs as a reference standard, newer methods exist for diagnosis of the disease. Serological assays for antibodies against SARS-CoV 2 are increasingly available and may represent a better gold standard in diagnosis for future research [37]. RT-PCR is limited by swabbing technique for nasopharyngeal samples and the fact that the virus is more avid in the lower respiratory tract [38]. However, many patients may not seroconvert prior to death limiting this test to survivors only.

Point of care lung ultrasound is a new technique for diagnosis of COVID-19 which may mitigate many of the issues noted with the modalities discussed so far. It has no radiation, is fast, cheap and may be able to detect lower respiratory tract disease unlike nasopharyngeal swab. However, there is limited evidence beyond small case series on its diagnostic accuracy [39–41]. Further, like other ultrasound techniques accuracy will likely be operator dependent [42] and experience will need to be built up for robust results in evaluating suspected COVID-19.

Finally, much research has been conducted in the use of artificial intelligence techniques to correctly diagnose COVID-19 based on imaging [43–45]. These techniques would obviate capacity limitations in reporting imaging as well as eliminate inter-reporter variability. However, as with any supervised machine learning technique, large, generalisable datasets, with correctly pre-classified positive and negative cases (which in turn will depend on a truly accurate reference standard) are needed [46].

## Conclusion

Chest X-ray has poor sensitivity and specificity in diagnosing COVID-19 in the general population during the pandemic. CT scanning has demonstrated excellent sensitivity and should strongly be considered during the pandemic in the initial assessment of COVID-19. This needs to be balanced against the risk of excess radiation with CT, where capacity allows.

## Summary box

### What is already known on this topic

- Small observational studies, predominantly in China, have reported on imaging features in COVID-19 after a confirmed RT-PCR swab test
- These studies have shown limited sensitivity for chest X-ray, but excellent sensitivity for CT scans, it is not possible to calculate the specificity of these modalities as they only included patients with confirmed COVID-19, therefore it is not possible to assess their utility in the general population who may or may not have COVID-19
- Literature on this general population attending emergency departments and the accuracy of these imaging techniques is limited
- International guidelines including from the British Society of Thoracic Imaging and American College of Radiology do not recommend the use of CT in initial evaluation of suspected COVID-19, largely due to capacity concerns

### What this study adds

- This study shows that Chest x-ray has poor sensitivity and specificity in patients with suspected COVID-19 attending the emergency department, whilst CT has excellent sensitivity and is 29% more sensitive than CXR in our study cohort; there was also poor agreement between CT and CXR findings in COVID-19
- Patients with indeterminate imaging without classical distribution of COVID-19 should still be considered at high risk of having the disease
- Our data suggest that CT should be employed more widely as an initial investigation, where capacity allows and balanced against the risk of excess radiation exposure

## Data Availability

Anonymized data is available on reasonable request from the corresponding author.

https://aborakati.github.io/Imaging-in-COVID/

## Acknowledgements

We would like to thank Scott Wilson from the Royal Free Hospital’s clinical practice group analytics department for retrieving the data from the hospital’s data warehouse.

We would like to thank Dr Federico Ricciardi of the Department of Statistical Science and PRIMENT Clinical Trials Unit at University College London for reviewing the statistical methods in this study.

## Data availability

Anonymised data is available on reasonable request from the corresponding author. Analysis scripts are attached as a supplementary file.

## Supplementary file 1

**Supplementary figure 1.**
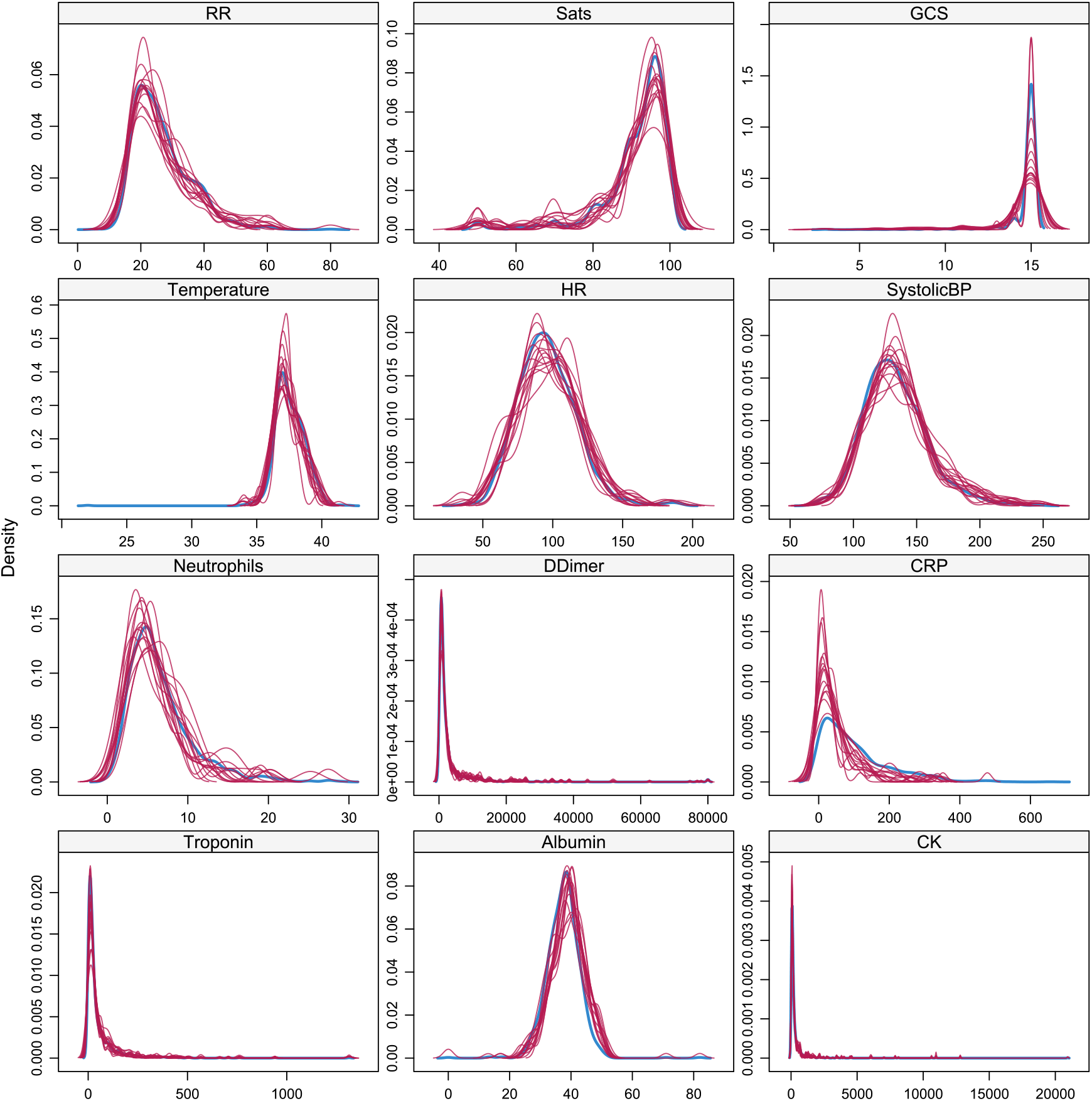
Density plots of imputed datasets; Blue represents original dataset; other colours are individual imputed datasets (n=15)

**Supplementary table 1.** Means of data before multiple imputation and propensity score matching.

| Covariate: | Means Treated | Means Control | Standard Deviation Control | Mean Difference |
| --- | --- | --- | --- | --- |
| **Overall Propensity Score** | 0.422997940 | 0.53935303 | 0.1449627 | -0.1163550897 |
| **Female** | 36.3782051 | 45.026178 | 0.4979547 | -8.64797288 |
| **Male** | 63.6217949 | 54.973822 | 0.4979547 | 8.64797288 |
| **Age** | 63.796474359 | 66.19022688 | 18.5893357 | -23.937525171 |
| **Comorbidity- Yes** | 76.1217949 | 84.467714 | 0.3625287 | -8.34591892 |
| **Ethnicity- South Asian** | 6.5705128 | 6.631763 | 0.2490539 | -0.06124983 |
| **Ethnicity- Black** | 16.1858974 | 11.518325 | 0.3195219 | 4.66757283 |
| **Ethnicity- Mixed** | 0.9615385 | 1.396161 | 0.1174340 | -0.43462210 |
| **Ethnicity- Other** | 18.9102564 | 13.263525 | 0.3394765 | 5.64673110 |
| **Ethnicity- White** | 46.6346154 | 57.766143 | 0.4943635 | -11.13152772 |
| **Respiratory Rate** | 29.214743590 | 24.01745201 | 7.2639816 | 5.1972915828 |

**Supplementary table 2.** Balance summary across imputations.

|  | Type | Minimum Difference Adjusted | Mean Difference Adjusted | Maximum Difference Adjusted |
| --- | --- | --- | --- | --- |
| **Distance** | Distance | 0.016988 | 0.027107 | 0.040963 |
| **Sex = Male** | Binary | -0.03917 | -0.0028 | 0.015982 |
| **Age** | Contin. | -0.04586 | -0.01371 | 0.027589 |
| **Comorbidity = Yes** | Binary | -0.02331 | -0.00778 | 0.004598 |
| **Ethnicity = Other Asian** | Binary | -0.01392 | 0.002362 | 0.016471 |
| **Ethnicity = South Asian** | Binary | -0.01399 | -0.00136 | 0.011905 |
| **Ethnicity = Black** | Binary | -0.01852 | 0.000443 | 0.015982 |
| **Ethnicity = Mixed** | Binary | -0.00464 | 0.001403 | 0.007042 |
| **Ethnicity = Other** | Binary | -0.01152 | 4.30E-06 | 0.00939 |
| **Ethnicity = White** | Binary | -0.02353 | -0.00285 | 0.018433 |
| **Respiratory Rate** | Contin. | -0.06157 | -0.03478 | -0.00442 |

**Supplementary table 3.** Average Sample sizes pre- and post-matching across imputed data sets.

|  | XR- Negative | XR- Positive | Total |
| --- | --- | --- | --- |
| All | 573 | 625 | 1,198 |
| Matched | 430 | 430 | 860 |
| Unmatched | 143 | 195 | 338 |
| Discarded | 0 | 0 | 0 |

**Supplementary figure 2.**
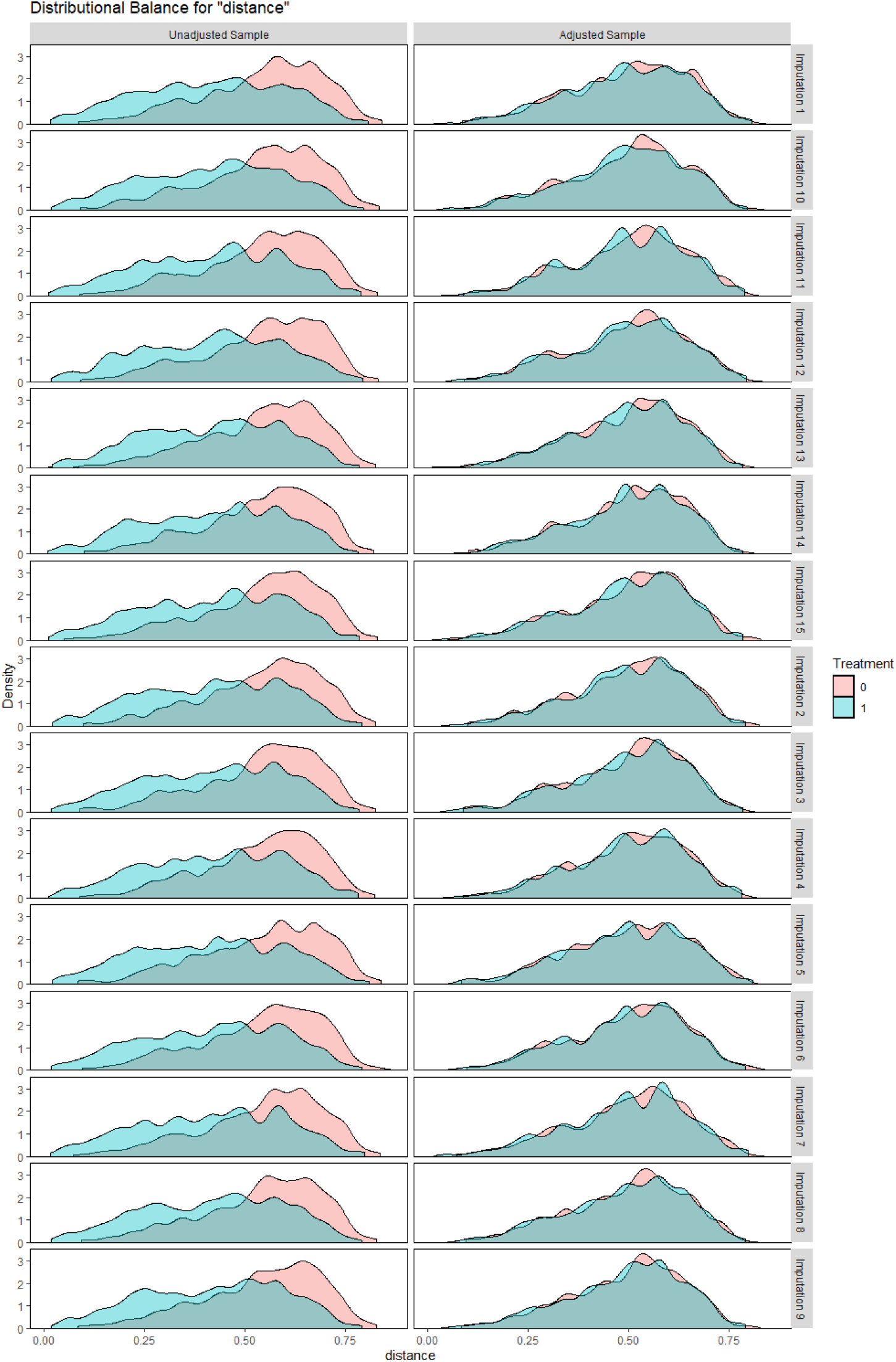
Density plot of propensity scores pre- and post-matching in each imputed dataset; treatment units re
present a positive X-ray for COVID-19, whereas a control unit represents a negative X-ray

**Supplementary figure 3.**
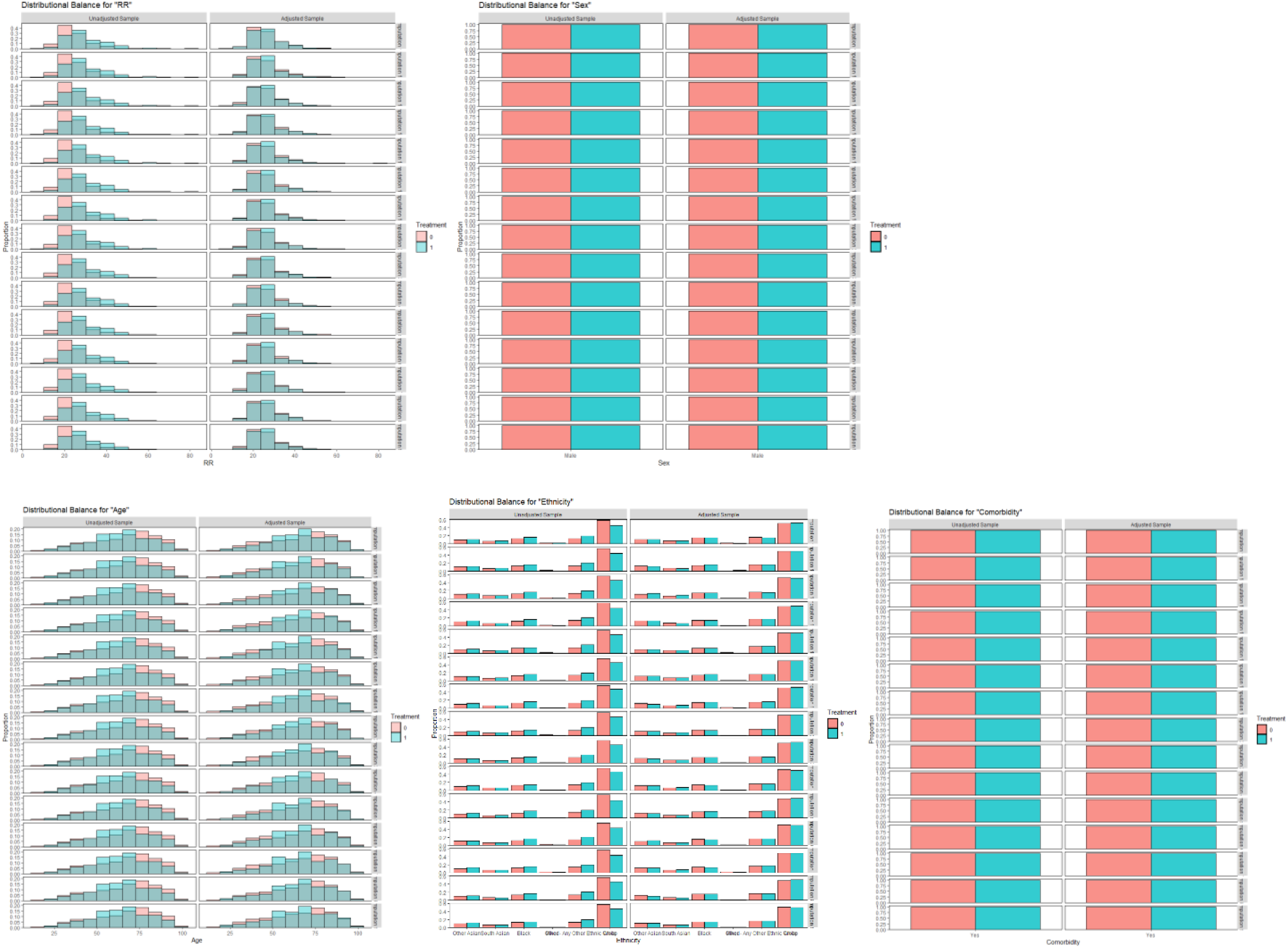
Histogram of distributions for each matching covariate pre- and post-matching in each imputed dataset; treatment units represent a positive X-ray for COVID-19, whereas a control unit represents a negative X-ray

**Supplementary figure 4.**
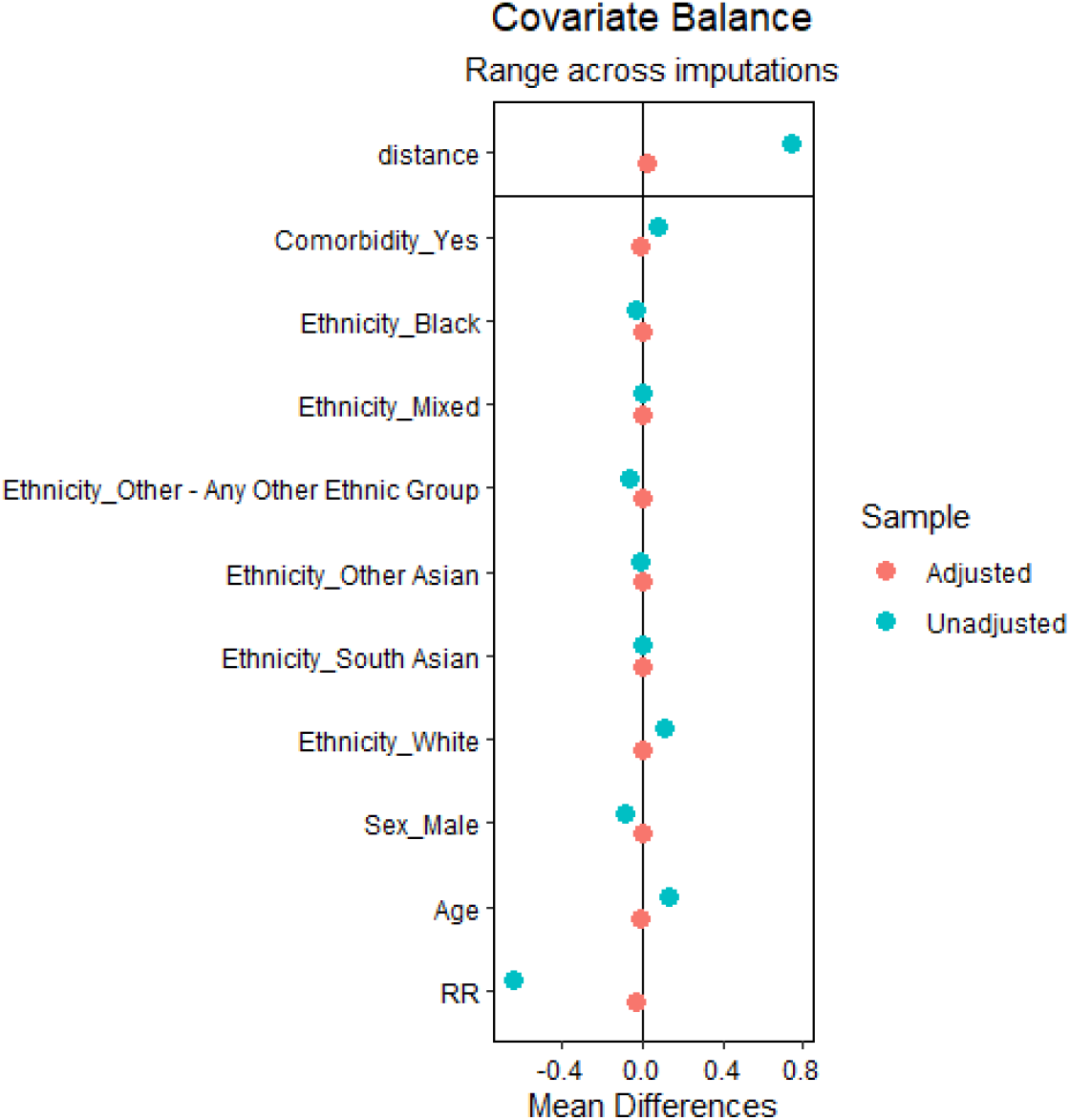
Love plot of pooled balances across imputed datasets in matching covariates after matching

